# Prioritizing COVID-19 tests based on participatory surveillance and spatial scanning

**DOI:** 10.1101/2020.05.25.20109058

**Authors:** OB Leal-Neto, FAS Santos, JY Lee, JO Albuquerque, WV Souza

## Abstract

**Objectives:** This study aimed to identify, describe and analyze priority areas for COVID-19 testing combining participatory surveillance and traditional surveillance.

**Design:** It was carried out a descriptive transversal study in the city of Caruaru, Pernambuco state, Brazil, within the period of 20/02/2020 to 05/05/2020. Data included all official reports for influenza-like illness notified by the municipality health department and the self-reports collected through the participatory surveillance platform Brasil Sem Corona.

**Methods:** We used linear regression and loess regression to verify a correlation between Participatory Surveillance (PS) and Traditional Surveillance (TS). Also a spatial scanning approach was deployed in order to identify risk clusters for COVID-19.

**Results:** In Caruaru, the PS had 861 active users, presenting an average of 1.2 reports per user per week. The platform Brasil Sem Corona started on March 20^th^ and since then, has been officially used by the Caruaru health authority to improve the quality of information from the traditional surveillance system. Regarding the respiratory syndrome cases from TS, 1,588 individuals were positive for this clinical outcome. The spatial scanning analysis detected 18 clusters and 6 of them presented statistical significance (p-value < 0.1). Clusters 3 and 4 presented an overlapping area that was chosen by the local authority to deploy the COVID-19 serology, where 50 individuals were tested. From there, 32% (n=16) presented reagent results for antibodies related to COVID-19.

**Conclusion:** Participatory surveillance is an effective epidemiological method to complement the traditional surveillance system in response to the COVID-19 pandemic by adding real-time spatial data to detect priority areas for COVID-19 testing.

## Introduction

Participatory surveillance has shown promising results from its conception to its application in several public health events [1-6]. The use of a collaborative information pathway provides a rapid way for the data collection on symptomatic individuals in the territory, to complement traditional health surveillance systems [7, 8]. In Brazil, this methodology has been used at the national level since 2014 during mass gatherings events since they have great importance for monitoring public health emergencies [9, 10]. With the occurrence of the COVID-19 pandemic, and the limitation of the main non-pharmaceutical interventions for epidemic control - in this case, testing and social isolation - added to the challenge of existing underreporting of cases and delay of notifications, there is a demand on alternative sources of up to date information to complement the current system for disease surveillance. Several studies [11-14] have demonstrated the benefits of participatory surveillance in coping with COVID-19, reinforcing the opportunity to modernize the way health surveillance has been carried out. Additionally, spatial scanning techniques have been used to understand syndromic scenarios, investigate outbreaks, and analyze epidemiological risk, constituting relevant tools for health management [15-18]. While there are limitations in the quality of traditional health systems, the data generated by participatory surveillance reveals an interesting application combining traditional techniques to clarify epidemiological risks that demand urgency in decision-making. Moreover, with the limitations of testing available, the identification of priority areas for intervention is an important activity in the early response to public health emergencies. This study aimed to describe and analyze priority areas for COVID-19 testing combining data from participatory surveillance and traditional surveillance for respiratory syndromes.

## Methods

The study’s method is a descriptive transversal, performed in the city of Caruaru, Pernambuco state, within the period of 02/20/2020 to 05/05/2020. Data included all official reports for influenza-like illness notified by municipality health department and the self-reports collected through the participatory surveillance platform Brasil Sem Corona[19].

To volunteer for the Brasil Sem Corona participatory surveillance platform, the individual should download an app called Colab. Through that, the volunteer has the option to agree with its terms of use that describe the purpose of this platform and ethical aspects, including the use of data for scientific studies that will help local public health authorities to improve the understanding of the epidemiological setting. After accepting the terms and conditions, participants can fill out a self-report questionnaire on symptoms and exposure in order to actively inform their health status. This questionnaire is based in a syndromic approach, including symptoms that are related with respiratory syndrome mixing up protocols used for participatory surveillance [9, 10]. Moreover, questions that could help to understand the severity of symptoms, such as if user sought a health facility or if it has elderly people as household members.

Daily reports are permitted, and a participant can report a maximum of two reports a day. Each time a participant completes and submits a report, it provides the respective latitude and longitude, which are anonymized in the server within a radius of 2 kilometers. Personal identification is also anonymized, and participants can drop out their enrollment in the platform at any time, and the data is completely deleted from the servers, following national privacy laws.

Data from traditional disease surveillance system was accessed by the epidemiological bulletins, publicly available at local government communication channels.

To verify a correlation between participatory surveillance (PS) and traditional surveillance (TS), it was carried out a linear regression [20] extracting summary of fit and parameter estimates:

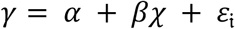

A local polynomial regression (LOESS) [21] was also performed to generalize a moving average to generate a scatterplot smoothing among the data points, where its function can be shown as:

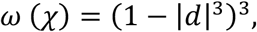

where *d* is the distance of data point from the point on the curve being fitted, scaled to lie in the range from 0 to 1.

On the spatial scanning method, it was performed a purely spatial analysis scanning for clusters with high rates using Bernoulli model assuming that in the studied area there are cases and non-cases represented by a dummy variable, acting as a spatial case (respiratory syndrome cases)/control (users that responded no symptoms) approach. It was decided not use temporal parameters for the SatScan analysis due to case-report delay from TS as well as the latency of 14 days that could be possible on COVID-19 cases.

The likelihood function for the Bernoulli [22] model is expressed as

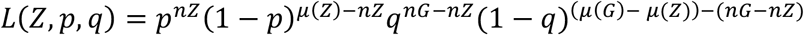

In order to detect the zone that is most likely to be a cluster, it was found the zone *Ž* that maximized the likelihood function, where *Ž* is the maximum likelihood estimator of the parameter *Z*. To achieve this, it needs to have two-stage actions. First, maximize the likelihood function conditioned on *Z*.

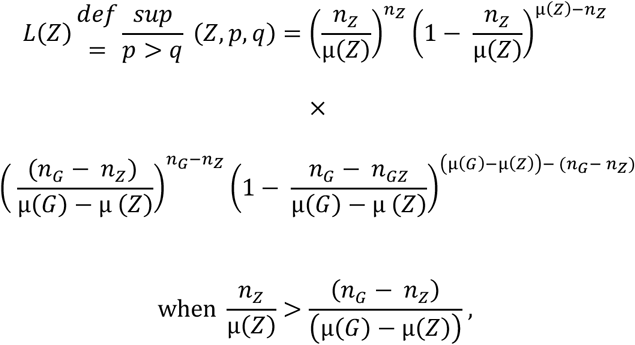

and otherwise

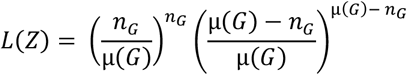

Second, to find the solution 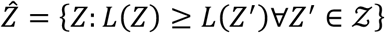.

The most likely cluster is of interest in itself, but it is important also to have a statistical inference. Let 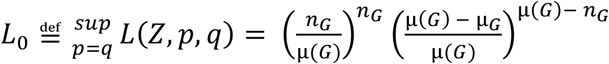

The likelihood ratio, *λ*, can be written as

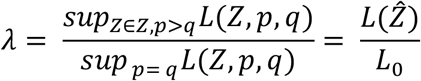

In addition, the SatScan runs a Monte Carlo simulation based on pseudorandom number generators. This allows to obtain the p-value for hypothesis testing, comparing the rank of the maximum likelihood from the real data set with the maximum likelihood from the random data sets.

In this case of spatial scanning clusters, statistical significance was defined as p-value < 0.1, bringing not only the clusters with a confidence interval of 95% and a maximum error of 5% but also considering a credible interval of 90% and a maximum error of 10%.

For the clusters that presented statistical significance [23], an overlaying in a geographic information system was done using QGIS 3.12 [24]. This map was used by the local authority to choose the area to deploy 50 serology tests for COVID-19. Other 50 tests were deployed in areas outside those clusters. For data wrangling, data tidying, and data visualization, an R language framework was used (Exploratory.io) [25]. For spatial scanning analysis, SatScan v9.6[26] was adopted.

The study had an exemption from the oversight body (Caruaru Health Department and Brasil Sem Corona scientific committee) since only public domain data available from secondary sources were used.

## Results

In Caruaru, the PS had 861 active users, presenting an average of 1.2 reports per user per week. The median age of users was 36 (mean 36.96; standard deviation 9.41; minimum age of 14 years old and maximum age of 60 years old).

The platform Brasil Sem Corona started on March 20^th^ 2020 and since then, has been officially used by the Caruaru health authority to improve the quality of information from the traditional surveillance system. Massive marketing campaigns using the official social media channels for the local government were used to advertise and engage new users.

Regarding the respiratory syndrome cases from TS, 1,588 individuals were positive for this clinical outcome. For the PS, it was considered 798 users that reported no symptoms in the period of study. Figures 1 and 2 shows the temporal and spatial distribution of cases (TS) and controls (PS).

**Figure 1.**
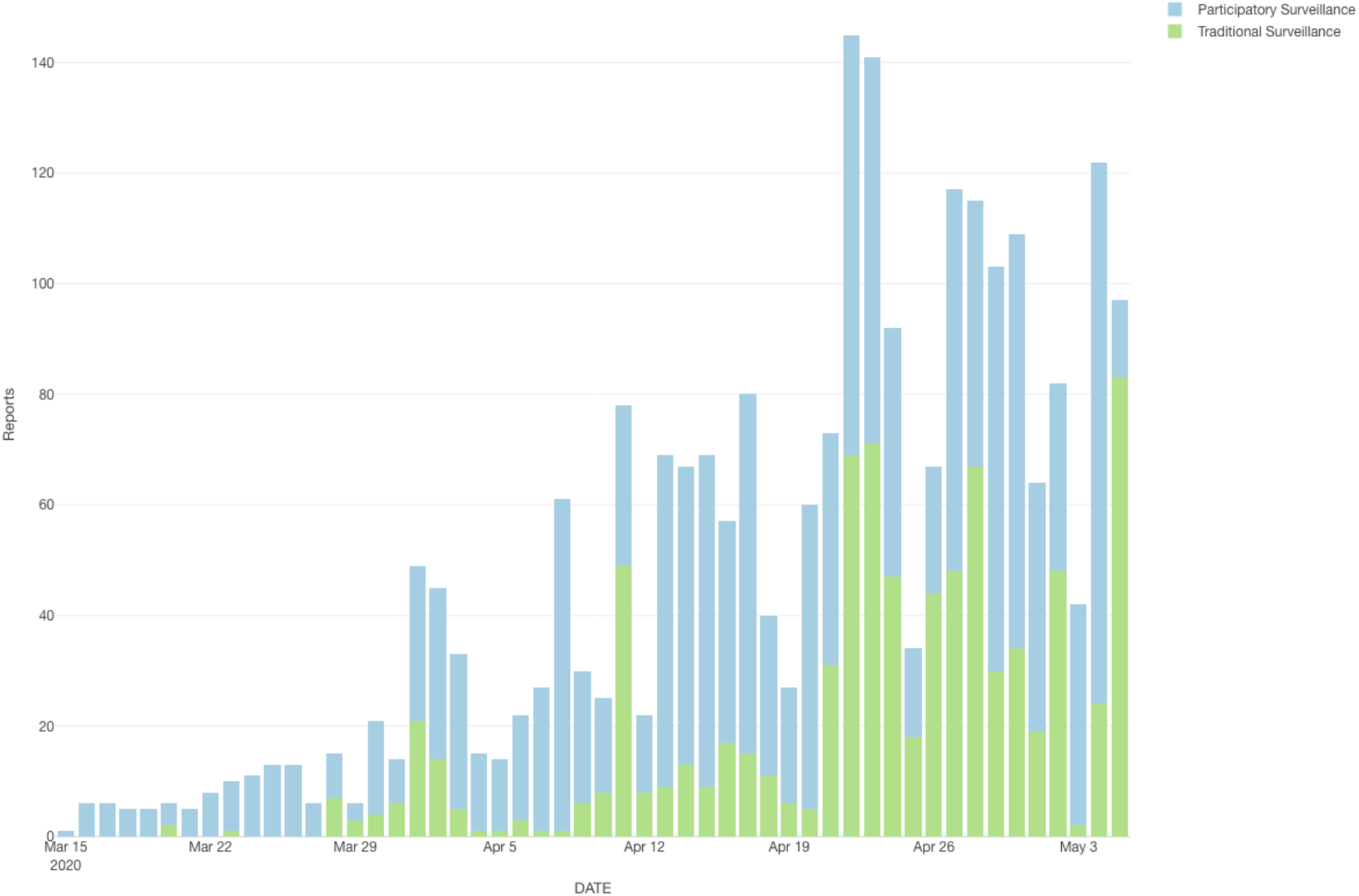
Temporal distribution of reports from traditional surveillance and participatory surveillance

**Figure 2.**
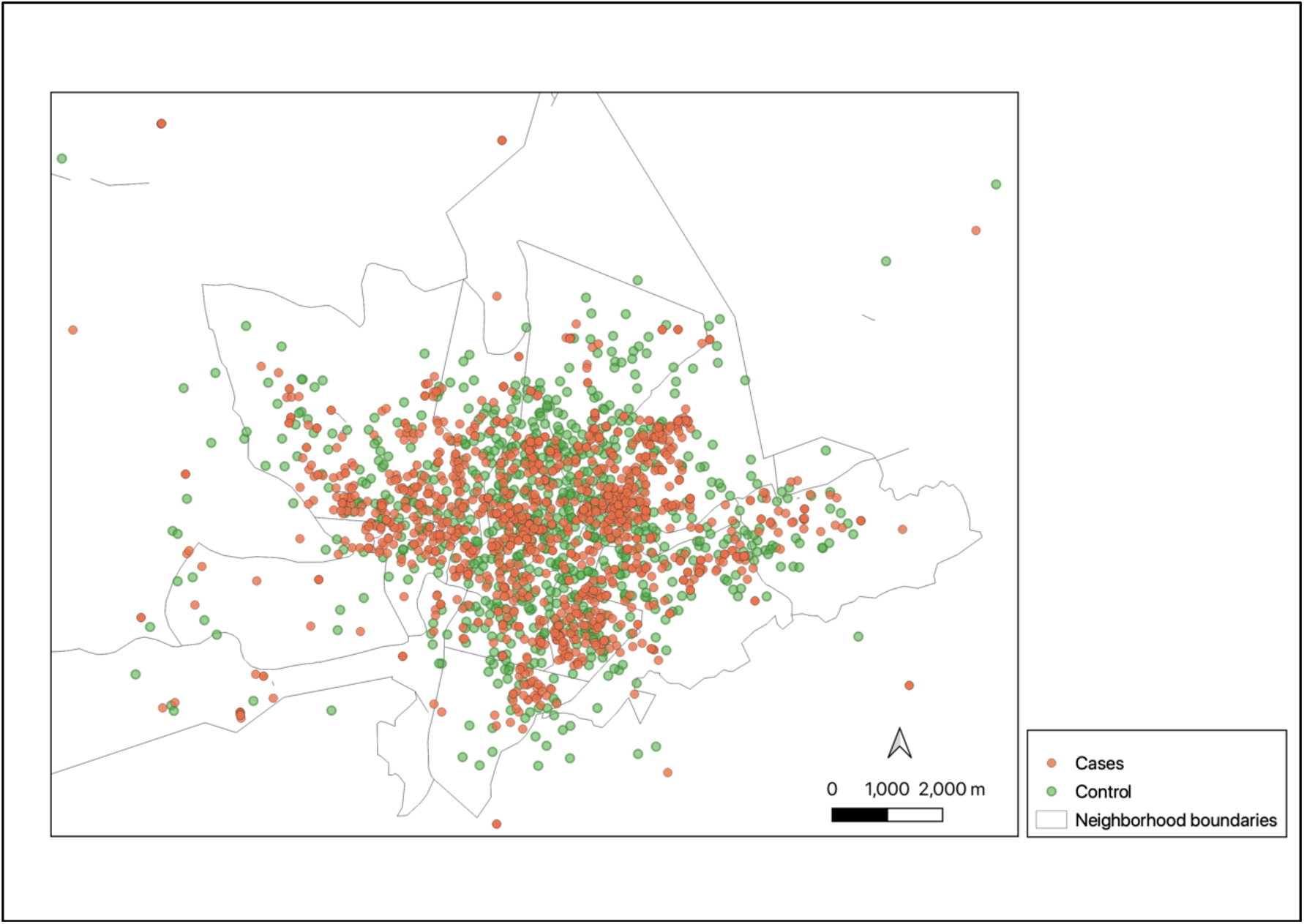
Spatial distribution of cases and controls over the city of Caruaru in the period of study.

When performed the signal (PS or TS) over time, it was found a linear correlation among these outputs and predictors where PS showed a correlation of 0.72 (R squared 0.52) and a coefficient of 1.17 (p-value < 0.001). When verified the results for TS, correlation showed an output of 0.6198 (R squared 0.38) and a coefficient of 1.13 (p-value < 0.001). Regarding the LOESS (Figure 3), PS has shown an equivalent number of parameters of 4.37, with a residual standard error of 17.00 and a trace of smoother matrix of 4.78. Finally, TS has shown as an equivalent number of parameters 4.62, residual standard error of 18.01, and a trace of smoother matrix of 5.07.

**Figure 3.**
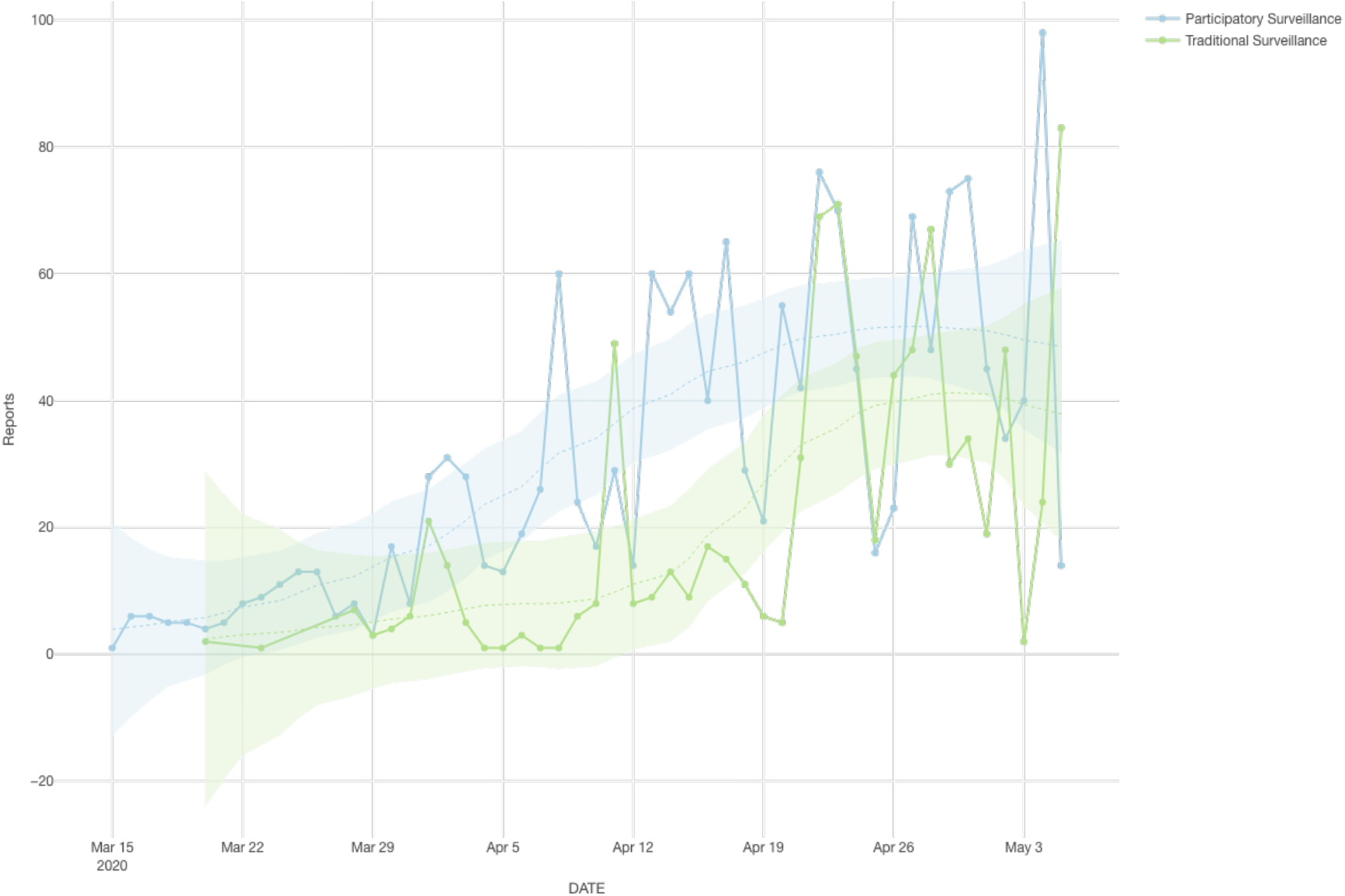
Time series distribution with LOESS regression for each source.

The spatial scanning analysis has detected 18 clusters and 6 of them have presented statistical significance (p-value < 0.1) (Table 1). Clusters 3 and 4 presented an overlapping and this area was chosen by the local authority to deploy the COVID-19 serology, where 50 individuals were tested (Figure 4). From there, 32% (*n*=16) presented reagent results for antibodies related to COVID-19, where the spatial scanning showed a hit rate between 84% and 94%. Another group of 50 individuals were tested outside identified clusters and from there, only 4% (n=2) were positive for COVID-19 serology.

**Table 1.** Summary of clusters including relative risk, log likelihood ratio and p-value.

| Cluster | Relative risk | Log likelihood ratio | P-value |
| --- | --- | --- | --- |
| 1 | 1.54 | 44.504679 | 0.001 |
| 2 | 1.53 | 26.568310 | 0.001 |
| 3 | 1.34 | 13.654698 | 0.003 |
| 4 | 1.38 | 12.143518 | 0.013 |
| 5 | 1.45 | 11.350351 | 0.021 |
| 6 | 1.46 | 9.770844 | 0.094 |
| 7 | 1.51 | 8.619205 | 0.274 |
| 8 | 1.37 | 8.025044 | 0.404 |
| 9 | 1.31 | 7.421699 | 0.544 |
| 10 | 1.51 | 7.382095 | 0.616 |
| 11 | 1.51 | 6.970157 | 0.728 |
| 12 | 1.51 | 6.970157 | 0.728 |
| 13 | 1.51 | 6.558434 | 0.848 |
| 14 | 1.35 | 6.492181 | 0.854 |
| 15 | 1.51 | 5.735634 | 0.975 |
| 16 | 1.29 | 5.413135 | 0.982 |
| 17 | 1.51 | 5.324557 | 0.994 |
| 18 | 1.51 | 5.324557 | 0.994 |

**Figure 4.**
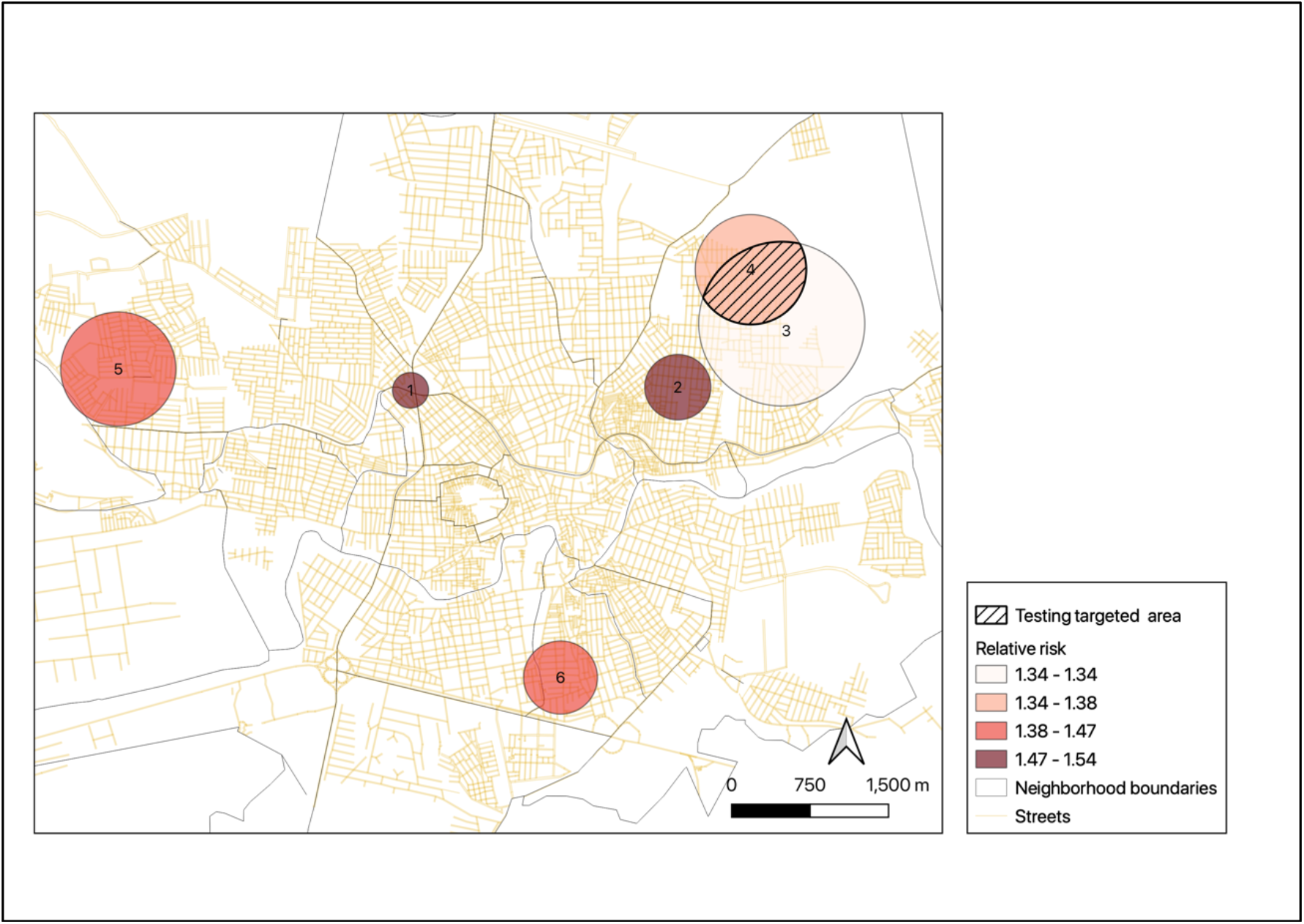
Caruaru city map showing the significant clusters, including an overlapping area between clusters 3 and 4 and testing targeted area.

## Discussion

The combination of TS and PS showed a relevant potential in the improvements of disease surveillance in the studied region, presenting 6 risk clusters that supported the decision of health managers. City areas that do not have health facilities can take advantage of the use of PS in order to cover blind spots for the TS, increasing the sensitivity and specificity of the official disease surveillance system.

There was a strong correlation between the participation of individuals in the PS strategy over time, whereas there was a moderate association in the same period with the increase in notifications related to influenza syndrome from TS.

Regarding spatial scanning, the use of clusters to target priority regions for testing was of relevant importance in the adequacy and rationalization of the use of diagnostic resources, which are sometimes scarce in different contexts. The finding of 32% people reagent for the COVID-19 antibody in areas that the analysis showed a variation between 34% and 38% demonstrates an excellent result for the method to be replicated not only in this city, but also in other places that need tools to prioritization of tests. The remaining clusters that did not overlap but were statistically significant will be implemented in the coming weeks.

The challenge of coping with COVID-19 is even greater in settings where there is a lack of access for testing, low schooling of the population, and minor investment in health policies. In addition, there are a large number of cases that are not registered due to the small number of symptoms [27] or by complete absence [28] of any symptoms that may guide accurate health care, at the right time and in the right place.

Brazil is one of the countries that have the lowest number of tests among those that have the highest number of confirmed cases for COVID-19, and one of the epicenters is the state of Pernambuco, where the number of cases can be up to 10 times greater than those officially presented in epidemiological reports [29]. The study region became known as one of the main areas affected by the Zika Virus syndrome in 2015 and 2016 [30], where there is a population that is affected by several problems such as measles, leprosy, and other diseases overcome elsewhere of the world. This increases the need for rapid response during the COVID-19 pandemic, since the health system is already overloaded with basal demands.

Even though most of the documented cases of COVID-19, in studies carried out in China, are in the age group between 30 to 79 years old [31], the fact we can observe in the PS a more prevalent age group of 36 is a relative trend on the use of digital mobile apps for a younger adults. This trend followed previous studies made in Brazil using participatory surveillance during mass gatherings [9,10]. In addition, for the present study another relevant finding is that were detected clusters were present in low-income areas that corroborate with other finds in Brazil [32].

In a study carried out by the Imperial College of London comparing the evolution of COVID-19 in 54 countries on several continents, it showed that Brazil is the country most threatened to be one of the main epicenters of the disease in the world, mainly due to the great possibility of spreading the disease with an R_0_ greater than 2 and an increasing number of deaths [33]. Knowledge of risk areas is essential for appropriate non-pharmacological interventions [34], as well as for the definition of priorities regarding medical care, for example. The best time to start treatment is another big challenge because as the disease has a rapid and almost silent evolution, knowing where the probable cases are can save lives, expand the possibility of differential diagnosis and favor an effective treatment.

The use of alternative methods as participatory surveillance showed a relevant role by taking advantage on the insertion at community levels, generating real-time spatial information not only for self-reported symptoms individuals but also for participants that informed to have no symptoms. Although the presented approach demands other sorts of validation and diagnostic methods (PCR) and it also needs to be carried out in different cities to address potential biases, our application shows an alternative to rapid mass screening in areas to prioritize tests and supporting local public health authorities to implement public policies based in evidence.

This research did not receive any specific grant from funding agencies in the public, commercial, or not-for-profit sectors.

## Data Availability

The data from Brasil Sem Corona can be obtained by formal requisition by www.brasilsemcorona.com.br
The data from respiratory syndrome was collected by the official disease surveillance bulletin provided by Caruaru Health Department, where is public available.

